# Core schema instability in youth at clinical high risk for psychosis: within-person variability as an index of self-belief precision

**DOI:** 10.64898/2026.10.27.26366553

**Authors:** Alexander Broekhuijse, Abhishek Saxena, Catalina Mourgues-Codern, Hesham Mukhtar, Jean Addington, Carrie E. Bearden, Kristin S. Cadenhead, Daniel H. Mathalon, Thomas H. McGlashan, Diana O. Perkins, Elaine F. Walker, Tyrone D. Cannon, Scott W. Woods, Emily A. Farina, Albert R. Powers

## Abstract

**Aim:** Core schemas are considered stable traits in clinical high risk (CHR) research. However, unexamined fluctuation may conceal belief changes that precede psychosis emergence. We examined whether schemas possess considerable within-person variability in CHR, and whether that variability exceeds that of controls.

**Methods:** In NAPLS-2, 411 CHR and 194 control participants completed the Brief Core Schema Scales (BCSS) at least three times across 24 months. Random-intercept models identified intraclass correlations in CHR. Bayesian censored location-scale models, correcting for floor scores and mean level differences, compared within-person SD between groups.

**Results:** Within-person variance accounted for 41.3%–50.6% of BCSS variance in CHR. Latent within-person SD was greater in CHR on all four subscales, most for negative-self (ratio 1.71, 95% CrI [1.50, 1.94]). This variability was largely independent of shared improvement and psychotropic medication effects.

**Conclusions:** Core schemas fluctuate substantially in CHR compared to controls, particularly in negative self-beliefs. Single baseline scores may understate schema–outcome associations.

## Introduction

The cognitive model of psychosis implicates core schemas—global and enduring beliefs about oneself and others (Jorovat et al., 2025)—in the development and interpretation of psychotic symptoms (Beck & Rector, 2005; Garety et al., 2001). Negative schemas about oneself and others have been proposed to orient the interpretation of anomalous social and sensory experiences toward persecutory and unusual interpretations (Freeman, 2007; Kesting & Lincoln, 2013). In youth at clinical high risk for psychosis (CHR), negative core schemas appear elevated relative to controls and have been examined as stable, trait-like predictors of symptoms and functioning (Cowan et al., 2019; Stowkowy et al., 2016). However, whether schemas actually behave as stable traits in this population has not been directly tested. This has implications for understanding psychotic symptom emergence, as a state-sensitive measure can capture progressive changes in beliefs and their relationship to other evolving symptoms that a trait-like measure cannot.

Several lines of work suggest that schemas may be unstable in CHR. Predictive processing accounts posit that the influence of new evidence on a belief depends on the precision, or certainty, attached to each (Adams et al., 2013; Sterzer et al., 2018). Paranoid ideation, for example, has been linked to expectations of volatility in one’s environment and the intentions of others (Hauke et al., 2024; Reed et al., 2020). Thus, beliefs about oneself and others held with low precision will likely shift with new experiences. Self-esteem, a narrower but associated construct, fluctuates significantly over hours and days, and this variability is linked to greater paranoia (Kesting & Lincoln, 2013; Thewissen et al., 2008, 2011). Whether schemas behave similarly in CHR is untested outside of small samples with two assessment points (Patton et al., 2022).

This study quantified, in a large CHR cohort, the degree to which core schemas fluctuate within individuals and whether this fluctuation differs from unaffected peers. We hypothesized that 1) schemas would fluctuate in CHR youth comparably to state-like measures, and 2) this fluctuation would be elevated relative to controls.

## Methods

### Design and sample

The sample comprised CHR participants and controls recruited through the second wave of the North American Prodrome Longitudinal Study (NAPLS-2) (Addington et al., 2012). CHR status was ascertained by the Structured Interview for Psychosis-Risk Syndromes, meeting Criteria of Prodromal Syndromes (Miller et al., 2003); controls were recruited in parallel. All participants completed written informed consent after receiving complete descriptions of the constituent studies, which were approved by local ethics committees. Assessments took place at baseline and at 6, 12, 18, and 24 months. This report included data from participants with at least three Brief Core Schema Scales (BCSS) administrations.

### Measures

The BCSS is a 24-item self-report measure of core beliefs about the self and others that is widely used in CHR (Stowkowy et al., 2016), forming four subscales (negative-self, negative-other, positive-self, and positive-other), each scored 0 to 24.

State-like benchmarks, all clinician-rated at the same visits, were the Scale of Prodromal Symptoms (SOPS) positive total (Miller et al., 2003), the Calgary Depression Scale for Schizophrenia (CDSS; Addington et al., 1993), and the Global Assessment of Functioning (GAF).

### Analysis

Analyses used R 4.5.1, with random-intercept models in lme4 1.1-37 (REML) and Bayesian censored location-scale models in brms 2.23.0 under default priors. For hypothesis 1 (schema fluctuation in CHR), an intercept-only model with a participant random intercept was fitted per subscale in CHR. Intraclass correlation coefficients (ICCs) identified the share of variance due to stable differences between people, and 1 − ICC the share due to change within people across visits, with 95% CIs from a 500-sample parametric bootstrap. Identical models were fitted to the SOPS positive total, CDSS, and GAF in the same CHR participants as state-like benchmarks. Each BCSS model was then refitted with visit as a categorical fixed effect to examine the amount of variance independent of general improvement.

Hypothesis 2 (greater fluctuation in CHR) was first tested by comparing intra-individual SDs between groups with Welch’s *t* tests and Cohen’s *d*, FDR-corrected across subscales. Because floor scores amongst controls make observed SDs appear artificially stable, the primary test used Bayesian censored (tobit) location-scale models, which accounted for floor and ceiling effects and estimated each group’s mean (location) and the spread of each person’s scores around their own mean (scale, or within-person SD). To account for group-based trajectory differences, group, months since baseline, their interaction, and a participant random intercept predicted the location; group predicted the scale. Fits used four chains of 4,000 iterations (2,000 warmup) and converged (R□ ≤ 1.01). The observed-SD comparison was repeated excluding the 15 CHR participants who developed psychosis during follow-up. The censored model was refitted with a single time trend for both groups, with visit as categorical, and with baseline age and psychotropic medication added.

## Results

### Sample characteristics

The final sample was 411 CHR participants (410 for negative-other) and 194 controls, with a mean of 4.06 and 4.18 assessments, respectively. Sample characteristics are outlined in **Table S1**. CHR participants were younger than controls, *t*(348.0) = 2.70, *p* = .007, and the groups did not differ in sex, χ^2^(1) = 2.23, *p* = .136. Participants retained and excluded by the three-assessment requirement did not differ in age, sex, or baseline schema level in either group (**Table S2**), but 37.7% of excluded CHR participants with follow-up data converted to psychosis compared with 3.6% of those retained.

### Core schemas demonstrate substantial within-person variance in CHR

Within-person variance accounted for about half of total BCSS variance in CHR: 49.0% (95% CI [44.3, 53.9]) for negative-self, 42.4% [38.3, 47.1] for negative-other, 41.3% [37.0, 45.8] for positive-self, and 50.6% [45.7, 55.3] for positive-other (**Table 1**; **Figure 1b**). Median intra-individual SDs ran from 2.35 to 3.09 points on the 0–24 scale, which on negative-self is approximately half the CHR sample’s own mean level of 4.85. To benchmark these values, the same model identified within-person variance of 55.1% for SOPS positive symptoms, 56.8% for GAF, and 63.2% for CDSS in these participants (**Table S4**).

**Table 1.**
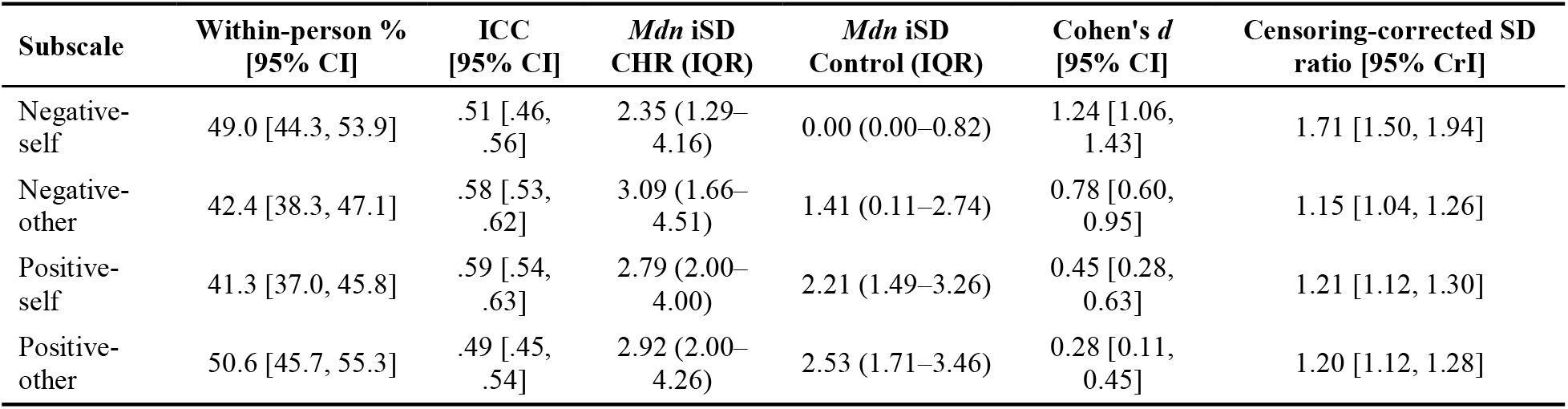
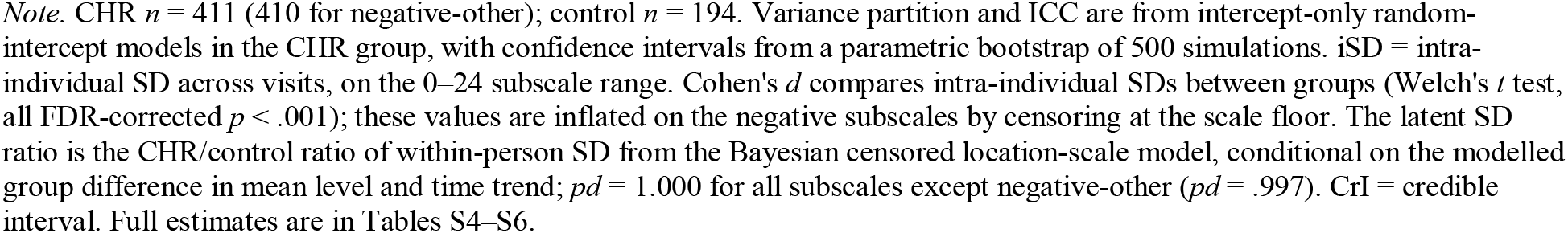
Within- and between-person variance in core schemas in CHR, and CHR–control comparison of within-person variability.

| Subscale | Within-person %<br>[95% CI] | ICC<br>[95% CI] | *Mdn* iSD<br>CHR (IQR) | *Mdn* iSD<br>Control (IQR) | Cohen's *d*<br>[95% CI] | Censoring-corrected SD<br>ratio [95% CrI] |
| --- | --- | --- | --- | --- | --- | --- |
| Negative-self | 49.0 [44.3, 53.9] | .51 [.46, .56] | 2.35 (1.29–4.16) | 0.00 (0.00–0.82) | 1.24 [1.06, 1.43] | 1.71 [1.50, 1.94] |
| Negative-other | 42.4 [38.3, 47.1] | .58 [.53, .62] | 3.09 (1.66–4.51) | 1.41 (0.11–2.74) | 0.78 [0.60, 0.95] | 1.15 [1.04, 1.26] |
| Positive-self | 41.3 [37.0, 45.8] | .59 [.54, .63] | 2.79 (2.00–4.00) | 2.21 (1.49–3.26) | 0.45 [0.28, 0.63] | 1.21 [1.12, 1.30] |
| Positive-other | 50.6 [45.7, 55.3] | .49 [.45, .54] | 2.92 (2.00–4.26) | 2.53 (1.71–3.46) | 0.28 [0.11, 0.45] | 1.20 [1.12, 1.28] |

Note. CHR $n = 411$ (410 for negative-other); control $n = 194$ . Variance partition and ICC are from intercept-only random-intercept models in the CHR group, with confidence intervals from a parametric bootstrap of 500 simulations. iSD = intra-individual SD across visits, on the 0–24 subscale range. Cohen's $d$ compares intra-individual SDs between groups (Welch's $t$ test, all FDR-corrected $p < .001$ ); these values are inflated on the negative subscales by censoring at the scale floor. The latent SD ratio is the CHR/control ratio of within-person SD from the Bayesian censored location-scale model, conditional on the modelled group difference in mean level and time trend; $pd = 1.000$ for all subscales except negative-other ( $pd = .997$ ). CrI = credible interval. Full estimates are in Tables S4–S6.

**Figure 1.**
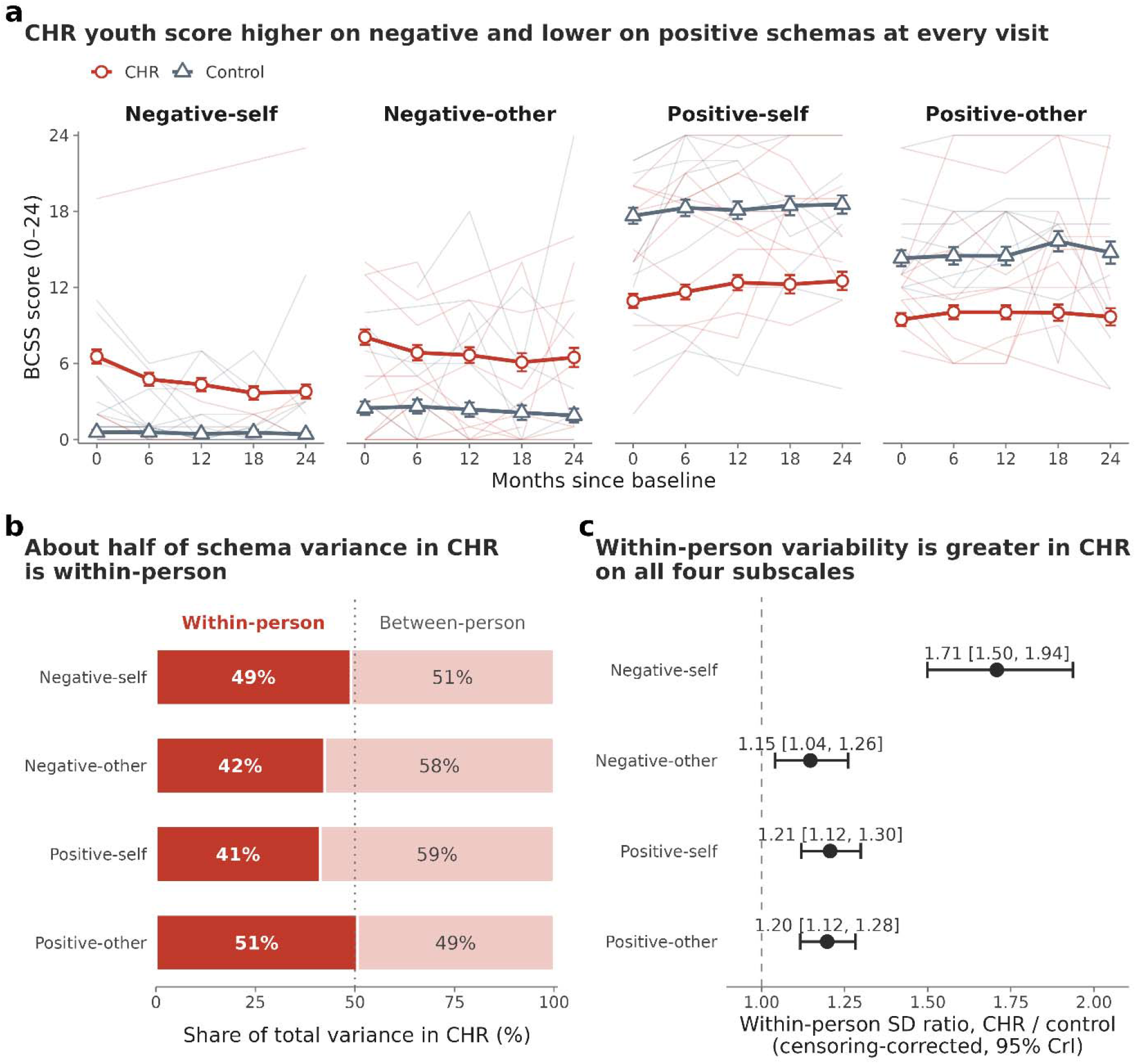
Core schemas fluctuate within CHR youth, and more than in controls. (a) Mean BCSS score by visit and group, with 95% confidence intervals; each mean uses all participants assessed at that visit (per-visit *n* in Table S3). Faint lines show 10 randomly selected participants per group. (b) Within- and between-person shares of total BCSS variance in CHR, from intercept-only random-intercept models; the dotted line marks 50%. (c) CHR/control ratio of within-person SD from Bayesian censored location-scale models with separate time trends by group, with 95% credible intervals; the dashed line marks equal variability.

### Within-person variability is greater in CHR than in controls on all four subscales

Intra-individual SDs were larger in CHR than in controls on every subscale (*d* = 0.28–1.24, all FDR-corrected *p* < .001; **Table S6**; **Figure S1**). The censoring-corrected model gave within-person SD ratios of 1.71 (95% CrI [1.50, 1.94]), 1.15 [1.04, 1.26], 1.21 [1.12, 1.30], and 1.20 [1.12, 1.28] for negative-self, negative-other, positive-self, and positive-other, respectively (**Figure 1c**; **Table S5**), indicating notably greater variability in core schemas amongst CHR youth compared to controls even when accounting for floor and ceiling effects and adjusting for mean schema levels. Refitting with a single shared time trend (negative-self 1.57 [1.38, 1.78]), categorical visit (1.67 [1.46, 1.89]), psychotropic medication use (1.67 [1.45, 1.90]), and baseline age (1.71 [1.49, 1.94]) left conclusions unchanged; other subscales moved by 0.08 or less (**Table S5**). Restricting the observed-SD comparison to the 396 non-converters left effect sizes unchanged (**Table S6**).

Adding visit as a fixed effect reduced residual within-person variance by only 0.2% to 10.0% across subscales (**Figure 1a**; **Table S4**), suggesting shared improvement accounted for little fluctuation.

## Discussion

Core schemas varied more within CHR youth compared to controls on all four subscales after accounting for the scale floor. The group difference was greatest in negative-self beliefs, where variability was 1.7 times higher. Schemas were nearly as variable as symptoms and functioning in the same participants, and far more variable than personality traits (17% within-person variance; Anusic & Schimmack, 2016).

Given that half of BCSS variance in CHR was within-person, a single administration may be a poor approximation of a participant’s schema level. Correlations between baseline schema scores and later symptoms, functioning, or conversion to psychosis may therefore underestimate effects. Averaging levels across visits will provide a more accurate estimate of a participant’s true schema level, and schema change itself may have clinical implications (Patton et al., 2022).

These findings are consistent with a developmental account of psychosis that emphasizes early belief change. Attenuated delusions often emerge before attenuated hallucinations in CHR and first-episode samples (Mourgues-Codern et al., 2025), and computational accounts propose that perceived volatility and negative affect mutually reinforce one another, overweighting negatively valenced priors (Benrimoh et al., 2025). Unstable schemas therefore may reflect this process, when beliefs about oneself and others are negative but not held with high precision. However, whether schema instability reflects an emerging illness or is itself a preexisting vulnerability cannot be settled in an already-symptomatic sample. Further, assessments separated by six months fail to capture the timescale social inference operates on. Future evaluations of belief change in CHR youth should use daily sampling of self-beliefs (Myin-Germeys et al., 2018), or design tasks manipulating self-relevant beliefs within a session which can estimate precision as a model parameter.

Two limitations qualify these findings. First, requiring three assessments removed participants who converted at roughly ten times the rate of those retained, so this is a CHR sample that largely did not transition; the converter sensitivity analysis is reassuring but underpowered. Second, variance in a self-report measure across visits conflates change in the underlying belief with measurement error. However, BCSS test-retest correlations imply that measurement error accounts for at most about 16% of score variance between occasions (Fowler et al., 2006), well below the 41–51% within-person share observed here, so most within-person variance in CHR cannot be attributed to measurement error.

## Conclusion

Core schemas are not purely traits in CHR youth. Instead, they fluctuate substantially, comparably to symptoms in the same participants, and more than in unaffected peers. The greatest group difference appeared in negative beliefs about the self. Therefore, schema change and variability may offer a more accurate assessment of self-beliefs than baseline level alone.

## Data Availability

All data produced in the present study are available upon reasonable request to the authors

## Acknowledgements

We gratefully acknowledge the contributions of Dr. Barbara A. Cornblatt, Dr. Lawrence J. Seidman, and Dr. Ming T. Tsuang, who each contributed substantially to the conception, design, and acquisition of data for the NAPLS consortium, meaningfully expanded the field of psychosis research, and who are now deceased. The work described in this article was funded in part by the State of Connecticut, Department of Mental Health and Addiction Services, but this publication does not express the views of the Department of Mental Health and Addiction Services or the State of Connecticut. The views and opinions expressed are those of the authors.

