## Supplementary material for "Core schema instability in youth at clinical high risk for psychosis: within-person variability as an index of self-belief precision": Online Supplement

*Table S1.* Sample characteristics by group

| Characteristic | CHR | Control | Test | *p* |
| --- | --- | --- | --- | --- |
| *n* | 411 | 194 |  |  |
| Age, *M* (*SD*) | 18.5 (4.36) | 19.6 (4.79) | *t*(348.0) = 2.70 | .007 |
| Female, *n* (%) | 169 (41.1%) | 93 (47.9%) | χ²(1) = 2.23 | .136 |
| *Race/ethnicity, n (%)* |  |  |  |  |
| Hispanic/Latino (any race) | 75 (18.2%) | 35 (18.0%) |  |  |
| Non-Hispanic American Indian/First Nations | 4 (1.0%) | 0 (0.0%) |  |  |
| Non-Hispanic Asian | 31 (7.5%) | 17 (8.8%) |  |  |
| Non-Hispanic Black/African American | 68 (16.5%) | 37 (19.1%) |  |  |
| Non-Hispanic Native Hawaiian/Pacific Islander | 1 (0.2%) | 1 (0.5%) |  |  |
| Non-Hispanic Other/Multiracial | 29 (7.1%) | 8 (4.1%) |  |  |
| Non-Hispanic White | 203 (49.4%) | 96 (49.5%) |  |  |
| Years of education, *M* (*SD*) | 11.33 (2.81) | 12.64 (3.62) |  |  |
| BCSS assessments, *M* (*SD*) | 4.06 (0.84) | 4.18 (0.77) |  |  |
| 3 assessments, *n* (%) | 134 (32.6%) | 43 (22.2%) |  |  |
| 4 assessments, *n* (%) | 117 (28.5%) | 74 (38.1%) |  |  |
| 5 assessments, *n* (%) | 160 (38.9%) | 77 (39.7%) |  |  |
| *Person-mean BCSS, M (SD)* |  |  |  |  |
| Negative-self | 4.85 (4.17) | 0.51 (0.81) |  |  |
| Negative-other | 6.90 (5.00) | 2.29 (2.84) |  |  |
| Positive-self | 11.83 (4.88) | 18.16 (3.72) |  |  |
| Positive-other | 9.83 (4.21) | 14.66 (3.81) |  |  |
| *Baseline clinical ratings, M (SD)* |  |  |  |  |
| SOPS positive total | 11.55 (3.91) | 1.13 (1.82) |  |  |
| CDSS total | 5.89 (4.84) | 0.65 (1.60) |  |  |
| GAF | 49.27 (11.17) | 83.84 (9.83) |  |  |
| Converted to psychosis, *n* (%) | 15 (3.6%) | — |  |  |

*Note.* Analytic sample: NAPLS-2 participants with at least three BCSS scores on at least one subscale (CHR *n* = 411; control *n* = 194). Age was compared with Welch's *t* test and sex with a χ² test with Yates' continuity correction; other rows are descriptive. BCSS assessments are the maximum number of scored visits across subscales. Person-mean BCSS is each participant's mean across visits on the 0–24 subscale range. Baseline clinical ratings are from the baseline visit. BCSS = Brief Core Schema Scales; CDSS = Calgary Depression Scale for Schizophrenia; CHR = clinical high risk for psychosis; GAF = Global Assessment of Functioning; SOPS = Scale of Prodromal Symptoms.

*Table S2.* Participants retained and excluded by the three-assessment requirement

| Characteristic | Excluded | Retained | Test | *p* |
| --- | --- | --- | --- | --- |
| **CHR** |  |  |  |  |
| *n* | 304 | 411 |  |  |
| Age, *M* (*SD*) | 18.4 (4.11) | 18.5 (4.36) | *t*(672.9) = −0.43 | .669 |
| Female, *n* (%) | 136 (44.7%) | 169 (41.1%) | χ²(1) = 0.79 | .373 |
| *Baseline BCSS, M (SD)* |  |  |  |  |
| Negative-self | 6.34 (5.65) | 6.55 (5.57), *n* = 401 | *t*(626.6) = −0.49 | .625 |
| Negative-other | 7.58 (6.04) | 8.08 (6.15), *n* = 401 | *t*(635.6) = −1.07 | .283 |
| Positive-self | 11.36 (6.03) | 10.94 (5.61), *n* = 400 | *t*(604.7) = 0.94 | .348 |
| Positive-other | 10.09 (5.57) | 9.46 (5.25), *n* = 401 | *t*(607.8) = 1.49 | .137 |
| **Controls** |  |  |  |  |
| *n* | 78 | 194 |  |  |
| Age, *M* (*SD*) | 20.0 (4.35) | 19.6 (4.79) | *t*(155.9) = 0.53 | .596 |
| Female, *n* (%) | 42 (53.8%) | 93 (47.9%) | χ²(1) = 0.56 | .455 |
| *Baseline BCSS, M (SD)* |  |  |  |  |
| Negative-self | 0.88 (1.73) | 0.57 (1.28) | *t*(90.5) = 1.33 | .187 |
| Negative-other | 2.14 (3.23) | 2.48 (3.70) | *t*(127.4) = −0.71 | .478 |
| Positive-self | 16.97 (5.13) | 17.65 (4.47) | *t*(100.9) = −0.96 | .337 |
| Positive-other | 13.86 (5.45) | 14.31 (4.43) | *t*(96.0) = −0.60 | .553 |

*Note.* Among NAPLS-2 participants with at least one BCSS score, participants were retained if they had at least three scores on at least one subscale. Age and baseline BCSS were compared with Welch's *t* tests and sex with a χ² test with Yates' continuity correction. Baseline BCSS is the score at the baseline visit; *n* is the number with a baseline score, which excludes some retained participants who lacked one. BCSS = Brief Core Schema Scales; CHR = clinical high risk for psychosis.

*Table S3.* BCSS coverage and floor and ceiling rates

|  | A. Participants with a score, *n* | | | | | B. Floor, ceiling, and variability | | | |
| --- | --- | --- | --- | --- | --- | --- | --- | --- | --- |
| Subscale | BL | 6 | 12 | 18 | 24 | Obs. at 0, % | Obs. at 24, % | All visits at 0, % of participants | *Mdn* iSD |
| **CHR** |  |  |  |  |  |  |  |  |  |
| Negative-self | 401 | 374 | 383 | 253 | 256 | 23.0 | 0.1 | 5.6 | 2.35 |
| Negative-other | 400 | 373 | 382 | 252 | 256 | 18.4 | 2.2 | 6.1 | 3.09 |
| Positive-self | 400 | 376 | 383 | 253 | 255 | 2.0 | 3.6 | 0.0 | 2.79 |
| Positive-other | 401 | 375 | 384 | 252 | 255 | 2.7 | 2.0 | 0.5 | 2.92 |
| **Controls** |  |  |  |  |  |  |  |  |  |
| Negative-self | 193 | 162 | 180 | 136 | 139 | 73.6 | 0.0 | 50.5 | 0.00 |
| Negative-other | 193 | 162 | 180 | 136 | 139 | 53.0 | 0.1 | 25.3 | 1.41 |
| Positive-self | 193 | 162 | 180 | 136 | 139 | 0.2 | 17.3 | 0.0 | 2.21 |
| Positive-other | 193 | 162 | 180 | 136 | 139 | 0.1 | 5.6 | 0.0 | 2.53 |

*Note.* Analytic sample (participants with at least three scores on the subscale). Panel A gives the number of participants with a score at baseline (BL) and at 6, 12, 18, and 24 months. Panel B gives the percentage of observations at the scale floor (0; left-censored in the location-scale models) and ceiling (24; right-censored), the percentage of participants who scored 0 at every visit, and the median intra-individual SD. BCSS = Brief Core Schema Scales; CHR = clinical high risk for psychosis; iSD = intra-individual standard deviation.

*Table S4.* Variance components in the CHR group

| Measure | *n* persons | *n* obs | τ_00_ | σ² | ICC [95% CI] | Within-person % [95% CI] | σ² with visit | Reduction in σ², % |
| --- | --- | --- | --- | --- | --- | --- | --- | --- |
| Negative-self | 411 | 1,667 | 13.69 | 13.16 | .51 [.46, .56] | 49.0 [44.3, 53.9] | 11.85 | 10.0 |
| Negative-other | 410 | 1,663 | 21.12 | 15.56 | .58 [.53, .62] | 42.4 [38.3, 47.1] | 14.93 | 4.0 |
| Positive-self | 411 | 1,667 | 20.10 | 14.13 | .59 [.54, .63] | 41.3 [37.0, 45.8] | 13.74 | 2.7 |
| Positive-other | 411 | 1,667 | 14.06 | 14.42 | .49 [.45, .54] | 50.6 [45.7, 55.3] | 14.39 | 0.2 |
| SOPS positive total | 411 | 1,725 | 10.08 | 12.39 | .45 [.40, .50] | 55.1 [50.2, 60.3] | — | — |
| CDSS total | 411 | 1,717 | 6.66 | 11.41 | .37 [.31, .42] | 63.2 [58.2, 68.8] | — | — |
| GAF | 411 | 1,724 | 75.81 | 99.61 | .43 [.38, .48] | 56.8 [51.8, 61.5] | — | — |

*Note.* All rows use the 411 retained CHR participants. BCSS rows use the analytic sample. Benchmark rows use every scheduled visit at which the measure was non-missing. Estimates are from intercept-only random-intercept models (REML); ICC = τ_00_ / (τ_00_ + σ²), and within-person % = 100 × (1 − ICC). 95% CIs are percentile intervals from 500 parametric bootstrap samples. σ² with visit is the residual variance after adding visit as a categorical fixed effect; the reduction is relative to σ². BCSS = Brief Core Schema Scales; CDSS = Calgary Depression Scale for Schizophrenia; CHR = clinical high risk for psychosis; GAF = Global Assessment of Functioning; ICC = intraclass correlation coefficient; SOPS = Scale of Prodromal Symptoms; σ² = within-person (residual) variance; τ_00_ = between-person variance.

*Table S5.* Censored location-scale models

| Parameter | Negative-self | Negative-other | Positive-self | Positive-other |
| --- | --- | --- | --- | --- |
| **A. Primary model: separate linear time trends by group** |  |  |  |  |
| *n* persons (CHR/control) | 411/194 | 410/194 | 411/194 | 411/194 |
| *n* observations | 2,477 | 2,473 | 2,477 | 2,477 |
| Left-censored (0), % | 39.6 | 29.7 | 1.5 | 1.9 |
| Right-censored (24), % | 0.1 | 1.5 | 8.1 | 3.2 |
| *Location* |  |  |  |  |
| Intercept | −2.97 [−3.84, −2.16] | 0.02 [−0.97, 0.97] | 18.18 [17.44, 18.92] | 14.40 [13.75, 15.06] |
| CHR vs control (at baseline) | 8.43 [7.44, 9.45] | 7.16 [6.03, 8.32] | −7.04 [−7.94, −6.15] | −4.70 [−5.54, −3.87] |
| Months since baseline, control | −0.027 [−0.059, 0.003] | −0.062 [−0.103, −0.022] | 0.044 [0.017, 0.072] | 0.036 [0.009, 0.065] |
| Months since baseline, CHR | −0.148 [−0.174, −0.122] | −0.098 [−0.128, −0.070] | 0.072 [0.048, 0.096] | 0.011 [−0.013, 0.035] |
| CHR × months | −0.121 [−0.161, −0.080] | −0.036 [−0.086, 0.012] | 0.028 [−0.009, 0.064] | −0.026 [−0.062, 0.012] |
| 24-month change, control | −0.66 [−1.41, +0.07] | −1.49 [−2.47, −0.52] | +1.05 [+0.41, +1.73] | +0.88 [+0.22, +1.55] |
| 24-month change, CHR | −3.56 [−4.18, −2.93] | −2.36 [−3.06, −1.69] | +1.72 [+1.16, +2.30] | +0.27 [−0.31, +0.84] |
| SD of participant intercepts | 4.31 [3.98, 4.67] | 5.57 [5.19, 6.01] | 4.54 [4.24, 4.87] | 3.85 [3.58, 4.13] |
| *Scale (log SD)* |  |  |  |  |
| Intercept | 0.89 [0.77, 1.01] | 1.38 [1.29, 1.46] | 1.17 [1.11, 1.24] | 1.20 [1.14, 1.26] |
| CHR | 0.54 [0.40, 0.66] | 0.14 [0.04, 0.23] | 0.19 [0.11, 0.26] | 0.18 [0.11, 0.25] |
| *Latent within-person SD* |  |  |  |  |
| Control | 2.43 [2.16, 2.75] | 3.96 [3.65, 4.32] | 3.23 [3.04, 3.44] | 3.31 [3.13, 3.51] |
| CHR | 4.15 [3.97, 4.34] | 4.54 [4.35, 4.75] | 3.90 [3.74, 4.06] | 3.96 [3.81, 4.13] |
| SD ratio (CHR/control) | 1.71 [1.50, 1.94] | 1.15 [1.04, 1.26] | 1.21 [1.12, 1.30] | 1.20 [1.12, 1.28] |
| *pd* | 1.000 | .997 | 1.000 | 1.000 |
| *Diagnostics* |  |  |  |  |
| Max R̂ | 1.004 | 1.004 | 1.004 | 1.006 |
| Min bulk ESS | 1,199 | 1,224 | 1,200 | 1,166 |
| Min tail ESS | 2,179 | 2,613 | 2,749 | 2,732 |
| **B. Panel A plus baseline age** |  |  |  |  |
| *n* persons (CHR/control) | 411/194 | 410/194 | 411/194 | 411/194 |
| *n* observations | 2,477 | 2,473 | 2,477 | 2,477 |
| Left-censored (0), % | 39.6 | 29.7 | 1.5 | 1.9 |
| Right-censored (24), % | 0.1 | 1.5 | 8.1 | 3.2 |
| *Location* |  |  |  |  |
| Intercept | −3.16 [−3.99, −2.32] | −0.01 [−1.00, 0.98] | 18.18 [17.43, 18.94] | 14.38 [13.71, 15.06] |
| CHR vs control (at baseline) | 8.69 [7.70, 9.67] | 7.18 [6.01, 8.42] | −7.03 [−7.95, −6.12] | −4.68 [−5.51, −3.85] |
| Months since baseline, control | −0.027 [−0.059, 0.004] | −0.061 [−0.103, −0.021] | 0.044 [0.016, 0.072] | 0.037 [0.009, 0.065] |
| Months since baseline, CHR | −0.148 [−0.175, −0.123] | −0.098 [−0.127, −0.069] | 0.072 [0.048, 0.096] | 0.011 [−0.012, 0.035] |
| CHR × months | −0.122 [−0.163, −0.080] | −0.037 [−0.085, 0.013] | 0.028 [−0.010, 0.065] | −0.025 [−0.062, 0.011] |
| 24-month change, control | −0.64 [−1.40, +0.10] | −1.47 [−2.47, −0.51] | +1.05 [+0.38, +1.73] | +0.88 [+0.21, +1.55] |
| 24-month change, CHR | −3.56 [−4.20, −2.94] | −2.35 [−3.05, −1.65] | +1.72 [+1.15, +2.30] | +0.27 [−0.30, +0.84] |
| Age (*z*) | 0.83 [0.44, 1.22] | 0.13 [−0.36, 0.63] | 0.08 [−0.31, 0.48] | 0.03 [−0.31, 0.38] |
| SD of participant intercepts | 4.25 [3.90, 4.62] | 5.57 [5.17, 6.01] | 4.54 [4.24, 4.86] | 3.84 [3.58, 4.12] |
| *Scale (log SD)* |  |  |  |  |
| Intercept | 0.89 [0.77, 1.01] | 1.38 [1.29, 1.46] | 1.17 [1.11, 1.24] | 1.20 [1.14, 1.26] |
| CHR | 0.54 [0.40, 0.66] | 0.14 [0.04, 0.23] | 0.19 [0.11, 0.26] | 0.18 [0.11, 0.25] |
| *Latent within-person SD* |  |  |  |  |
| Control | 2.43 [2.16, 2.75] | 3.96 [3.64, 4.33] | 3.23 [3.04, 3.44] | 3.31 [3.13, 3.51] |
| CHR | 4.15 [3.97, 4.34] | 4.55 [4.35, 4.75] | 3.90 [3.75, 4.06] | 3.96 [3.81, 4.13] |
| SD ratio (CHR/control) | 1.71 [1.49, 1.94] | 1.15 [1.04, 1.26] | 1.21 [1.12, 1.30] | 1.20 [1.11, 1.28] |
| *pd* | 1.000 | .997 | 1.000 | 1.000 |
| Change in SD ratio from Panel A | +0.000 | +0.000 | −0.001 | −0.001 |
| *Diagnostics* |  |  |  |  |
| Max R̂ | 1.003 | 1.003 | 1.004 | 1.003 |
| Min bulk ESS | 1,296 | 1,188 | 1,382 | 1,400 |
| Min tail ESS | 2,441 | 2,588 | 2,564 | 2,512 |
| **C. Single time trend shared by both groups** |  |  |  |  |
| *n* persons (CHR/control) | 411/194 | 410/194 | 411/194 | 411/194 |
| *n* observations | 2,477 | 2,473 | 2,477 | 2,477 |
| Left-censored (0), % | 39.6 | 29.7 | 1.5 | 1.9 |
| Right-censored (24), % | 0.1 | 1.5 | 8.1 | 3.2 |
| *Location* |  |  |  |  |
| Intercept | −2.42 [−3.24, −1.65] | 0.31 [−0.59, 1.23] | 18.02 [17.31, 18.73] | 14.54 [13.92, 15.16] |
| CHR vs control (at baseline) | 7.44 [6.53, 8.37] | 6.74 [5.70, 7.82] | −6.76 [−7.60, −5.93] | −4.96 [−5.68, −4.22] |
| Months since baseline | −0.103 [−0.126, −0.081] | −0.086 [−0.110, −0.063] | 0.060 [0.042, 0.078] | 0.022 [0.004, 0.040] |
| 24-month change (24 × slope) | −2.48 [−3.02, −1.94] | −2.07 [−2.65, −1.50] | +1.44 [+1.01, +1.88] | +0.53 [+0.10, +0.96] |
| SD of participant intercepts | 4.34 [4.00, 4.72] | 5.56 [5.16, 5.99] | 4.55 [4.24, 4.88] | 3.84 [3.58, 4.13] |
| *Scale (log SD)* |  |  |  |  |
| Intercept | 0.97 [0.85, 1.10] | 1.38 [1.30, 1.47] | 1.17 [1.11, 1.24] | 1.20 [1.14, 1.25] |
| CHR | 0.45 [0.32, 0.58] | 0.14 [0.03, 0.23] | 0.19 [0.11, 0.26] | 0.18 [0.11, 0.25] |
| *Latent within-person SD* |  |  |  |  |
| Control | 2.65 [2.35, 3.00] | 3.97 [3.65, 4.34] | 3.24 [3.04, 3.45] | 3.31 [3.13, 3.50] |
| CHR | 4.15 [3.97, 4.35] | 4.54 [4.35, 4.76] | 3.90 [3.75, 4.06] | 3.96 [3.81, 4.13] |
| SD ratio (CHR/control) | 1.57 [1.38, 1.78] | 1.15 [1.04, 1.26] | 1.20 [1.12, 1.30] | 1.20 [1.12, 1.28] |
| *pd* | 1.000 | .996 | 1.000 | 1.000 |
| Change in SD ratio from Panel A | −0.139 | −0.002 | −0.002 | +0.001 |
| *Diagnostics* |  |  |  |  |
| Max R̂ | 1.005 | 1.004 | 1.006 | 1.003 |
| Min bulk ESS | 1,485 | 1,238 | 850 | 1,387 |
| Min tail ESS | 2,873 | 2,521 | 1,858 | 2,733 |
| **D. Separate mean for each group at each visit** |  |  |  |  |
| *n* persons (CHR/control) | 411/194 | 410/194 | 411/194 | 411/194 |
| *n* observations | 2,477 | 2,473 | 2,477 | 2,477 |
| Left-censored (0), % | 39.6 | 29.7 | 1.5 | 1.9 |
| Right-censored (24), % | 0.1 | 1.5 | 8.1 | 3.2 |
| *Location* |  |  |  |  |
| Intercept | −2.98 [−3.88, −2.11] | −0.24 [−1.32, 0.82] | 18.05 [17.26, 18.85] | 14.39 [13.67, 15.11] |
| CHR vs control (at baseline) | 9.10 [8.02, 10.18] | 7.87 [6.62, 9.18] | −7.10 [−8.08, −6.09] | −4.94 [−5.85, −4.02] |
| Visit 24 vs baseline, control | −0.75 [−1.64, +0.09] | −1.39 [−2.53, −0.29] | +0.94 [+0.20, +1.71] | +0.43 [−0.32, +1.16] |
| Visit 24 vs baseline, CHR | −3.35 [−4.03, −2.64] | −2.13 [−2.89, −1.37] | +1.61 [+0.96, +2.24] | +0.18 [−0.47, +0.81] |
| SD of participant intercepts | 4.33 [4.00, 4.71] | 5.58 [5.20, 6.02] | 4.54 [4.24, 4.88] | 3.85 [3.59, 4.13] |
| *Scale (log SD)* |  |  |  |  |
| Intercept | 0.90 [0.78, 1.02] | 1.38 [1.29, 1.47] | 1.17 [1.11, 1.24] | 1.19 [1.13, 1.25] |
| CHR | 0.51 [0.38, 0.64] | 0.13 [0.03, 0.23] | 0.19 [0.11, 0.26] | 0.18 [0.11, 0.25] |
| *Latent within-person SD* |  |  |  |  |
| Control | 2.45 [2.19, 2.79] | 3.97 [3.65, 4.34] | 3.23 [3.04, 3.45] | 3.29 [3.11, 3.49] |
| CHR | 4.09 [3.91, 4.29] | 4.53 [4.34, 4.73] | 3.90 [3.74, 4.06] | 3.96 [3.80, 4.13] |
| SD ratio (CHR/control) | 1.67 [1.46, 1.89] | 1.14 [1.03, 1.26] | 1.21 [1.12, 1.30] | 1.20 [1.12, 1.29] |
| *pd* | 1.000 | .996 | 1.000 | 1.000 |
| Change in SD ratio from Panel A | −0.039 | −0.008 | +0.000 | +0.004 |
| *Diagnostics* |  |  |  |  |
| Max R̂ | 1.004 | 1.004 | 1.004 | 1.004 |
| Min bulk ESS | 1,217 | 1,071 | 901 | 1,250 |
| Min tail ESS | 2,200 | 2,095 | 1,940 | 2,209 |
| **E. Panel A plus time-varying psychotropic medication (location and scale)** |  |  |  |  |
| *n* persons (CHR/control) | 411/194 | 410/194 | 411/194 | 411/194 |
| *n* observations | 2,477 | 2,473 | 2,477 | 2,477 |
| Left-censored (0), % | 39.6 | 29.7 | 1.5 | 1.9 |
| Right-censored (24), % | 0.1 | 1.5 | 8.1 | 3.2 |
| *Location* |  |  |  |  |
| Intercept | −2.98 [−3.83, −2.14] | 0.00 [−1.02, 0.95] | 18.17 [17.44, 18.93] | 14.38 [13.75, 15.03] |
| CHR vs control (at baseline) | 8.34 [7.32, 9.41] | 7.22 [6.07, 8.50] | −6.80 [−7.76, −5.87] | −4.65 [−5.49, −3.80] |
| Months since baseline, control | −0.028 [−0.060, 0.003] | −0.063 [−0.103, −0.022] | 0.045 [0.017, 0.073] | 0.037 [0.010, 0.064] |
| Months since baseline, CHR | −0.146 [−0.172, −0.121] | −0.100 [−0.129, −0.071] | 0.072 [0.048, 0.095] | 0.012 [−0.012, 0.036] |
| CHR × months | −0.119 [−0.159, −0.078] | −0.037 [−0.087, 0.013] | 0.027 [−0.010, 0.063] | −0.025 [−0.061, 0.011] |
| 24-month change, control | −0.66 [−1.44, +0.07] | −1.51 [−2.48, −0.53] | +1.09 [+0.40, +1.76] | +0.89 [+0.23, +1.54] |
| 24-month change, CHR | −3.51 [−4.12, −2.90] | −2.40 [−3.10, −1.70] | +1.73 [+1.15, +2.29] | +0.29 [−0.29, +0.86] |
| Antipsychotic | 0.11 [−0.63, 0.88] | −1.33 [−2.25, −0.40] | −0.88 [−1.64, −0.13] | 0.18 [−0.55, 0.90] |
| Antidepressant | 0.43 [−0.29, 1.12] | 0.34 [−0.45, 1.14] | −0.41 [−1.07, 0.26] | −0.21 [−0.82, 0.42] |
| Other psychotropic | −0.32 [−1.08, 0.41] | 0.57 [−0.21, 1.45] | 0.23 [−0.45, 0.90] | −0.09 [−0.76, 0.59] |
| SD of participant intercepts | 4.32 [3.98, 4.69] | 5.58 [5.22, 6.02] | 4.53 [4.24, 4.85] | 3.86 [3.59, 4.13] |
| *Scale (log SD)* |  |  |  |  |
| Intercept | 0.88 [0.76, 1.00] | 1.38 [1.29, 1.46] | 1.17 [1.11, 1.23] | 1.20 [1.14, 1.25] |
| CHR | 0.51 [0.37, 0.64] | 0.11 [0.01, 0.21] | 0.13 [0.04, 0.20] | 0.15 [0.08, 0.23] |
| Antipsychotic | −0.20 [−0.33, −0.07] | 0.01 [−0.12, 0.15] | 0.06 [−0.05, 0.18] | 0.03 [−0.08, 0.15] |
| Antidepressant | 0.09 [−0.03, 0.21] | 0.08 [−0.03, 0.18] | 0.15 [0.05, 0.25] | −0.01 [−0.10, 0.10] |
| Other psychotropic | 0.19 [0.07, 0.31] | 0.01 [−0.10, 0.13] | 0.02 [−0.09, 0.13] | 0.10 [0.00, 0.20] |
| *Latent within-person SD (unmedicated visits)* |  |  |  |  |
| Control | 2.40 [2.13, 2.72] | 3.96 [3.64, 4.31] | 3.22 [3.03, 3.43] | 3.31 [3.13, 3.51] |
| CHR | 4.00 [3.78, 4.24] | 4.41 [4.17, 4.68] | 3.65 [3.46, 3.85] | 3.86 [3.67, 4.07] |
| SD ratio (CHR/control) | 1.67 [1.45, 1.90] | 1.11 [1.01, 1.24] | 1.13 [1.05, 1.23] | 1.17 [1.08, 1.26] |
| *pd* | 1.000 | .981 | .999 | 1.000 |
| Change in SD ratio from Panel A | −0.040 | −0.033 | −0.073 | −0.031 |
| *Diagnostics* |  |  |  |  |
| Max R̂ | 1.004 | 1.005 | 1.003 | 1.003 |
| Min bulk ESS | 1,254 | 690 | 1,287 | 1,311 |
| Min tail ESS | 2,264 | 1,410 | 2,519 | 2,994 |

*Note.* Bayesian censored (tobit) location-scale models fitted separately to each subscale in the analytic sample; scores of 0 and 24 were treated as left- and right-censored. Values are posterior medians [95% CrI]. Control is the reference group; CHR vs control is the location difference at baseline. Panel A is the primary model (location: group, months since baseline, their interaction, and a participant random intercept; scale: group); 24-month change is 24 times the group's monthly slope. Panel B adds standardised baseline age (*z*) to the location. Panel C fits a single time trend shared by both groups. Panel D replaces the linear time trend with a separate mean for each group at each visit; visit 24 vs baseline is the change in that group's mean. Panel E adds three time-varying indicators of medication current at each assessment (antipsychotic; antidepressant; other psychotropic, comprising mood stabilisers, stimulants, non-stimulant ADHD medication, anxiolytics, and hypnotics), coded from the NAPLS-2 prescription medication log, to both the location and the scale, so its latent within-person SDs and SD ratio are for unmedicated visits. Latent within-person SD is exp(scale intercept) for controls and exp(scale intercept + CHR) for CHR; the SD ratio is exp(CHR scale coefficient). *pd* is the probability of direction for the SD ratio. Change in SD ratio is the panel median minus the Panel A median. R̂ and effective sample sizes (ESS) are across all model parameters. ADHD = attention-deficit/hyperactivity disorder; CHR = clinical high risk for psychosis; CrI = credible interval; ESS = effective sample size.

*Table S6.* Observed intra-individual SD: group comparisons and non-converter sensitivity

|  | CHR | | | Control | | | Welch test | | | Effect size | | Mann–Whitney | | Non-converters | | |
| --- | --- | --- | --- | --- | --- | --- | --- | --- | --- | --- | --- | --- | --- | --- | --- | --- |
| Subscale | *n* | *M* (*SD*) | *Mdn* (IQR) | *n* | *M* (*SD*) | *Mdn* (IQR) | *t* (*df*) | *p* | *p*_FDR_ | *d* [95% CI] | *g* | *W* | *p* | CHR *n* | *d* [95% CI] | *p*_FDR_ |
| Negative-self | 411 | 2.91 (2.24) | 2.35 (1.29–4.16) | 194 | 0.55 (0.82) | 0.00 (0.00–0.82) | −18.88 (577.7) | < .001 | < .001 | 1.24 [1.06, 1.43] | 1.24 | 10,083 | < .001 | 396 | 1.24 [1.06, 1.43] | < .001 |
| Negative-other | 410 | 3.29 (2.15) | 3.09 (1.66–4.51) | 194 | 1.74 (1.65) | 1.41 (0.11–2.74) | −9.78 (481.5) | < .001 | < .001 | 0.78 [0.60, 0.95] | 0.78 | 22,194 | < .001 | 395 | 0.77 [0.60, 0.95] | < .001 |
| Positive-self | 411 | 3.22 (1.96) | 2.79 (2.00–4.00) | 194 | 2.39 (1.49) | 2.21 (1.49–3.26) | −5.73 (483.7) | < .001 | < .001 | 0.45 [0.28, 0.63] | 0.45 | 29,982 | < .001 | 396 | 0.45 [0.28, 0.62] | < .001 |
| Positive-other | 411 | 3.27 (1.89) | 2.92 (2.00–4.26) | 194 | 2.77 (1.68) | 2.53 (1.71–3.46) | −3.32 (423.2) | < .001 | < .001 | 0.28 [0.11, 0.45] | 0.28 | 33,496 | .001 | 396 | 0.28 [0.11, 0.45] | < .001 |

*Note.* Intra-individual SD (iSD) is each participant's SD across visits on the 0–24 subscale range, in the analytic sample. Welch's *t* compares control minus CHR. *p*_FDR_ is Benjamini–Hochberg adjusted across the four subscales. Cohen's *d* uses the pooled SD, with a normal-approximation 95% CI; *g* is Hedges' small-sample correction. *W* is the Mann–Whitney statistic. Non-converters: controls plus CHR participants with a conversion flag of 0 (all retained CHR participants had conversion follow-up data); *d* and *p*_FDR_ as above. Conversion is the authoritative NAPLS-2 conversion flag; participants without conversion follow-up data are excluded from conversion denominators. On the negative subscales, *t* and *d* are inflated by the scale floor. CHR = clinical high risk for psychosis; IQR = interquartile range.

*Figure S1.* Intra-individual SD of BCSS subscales by group

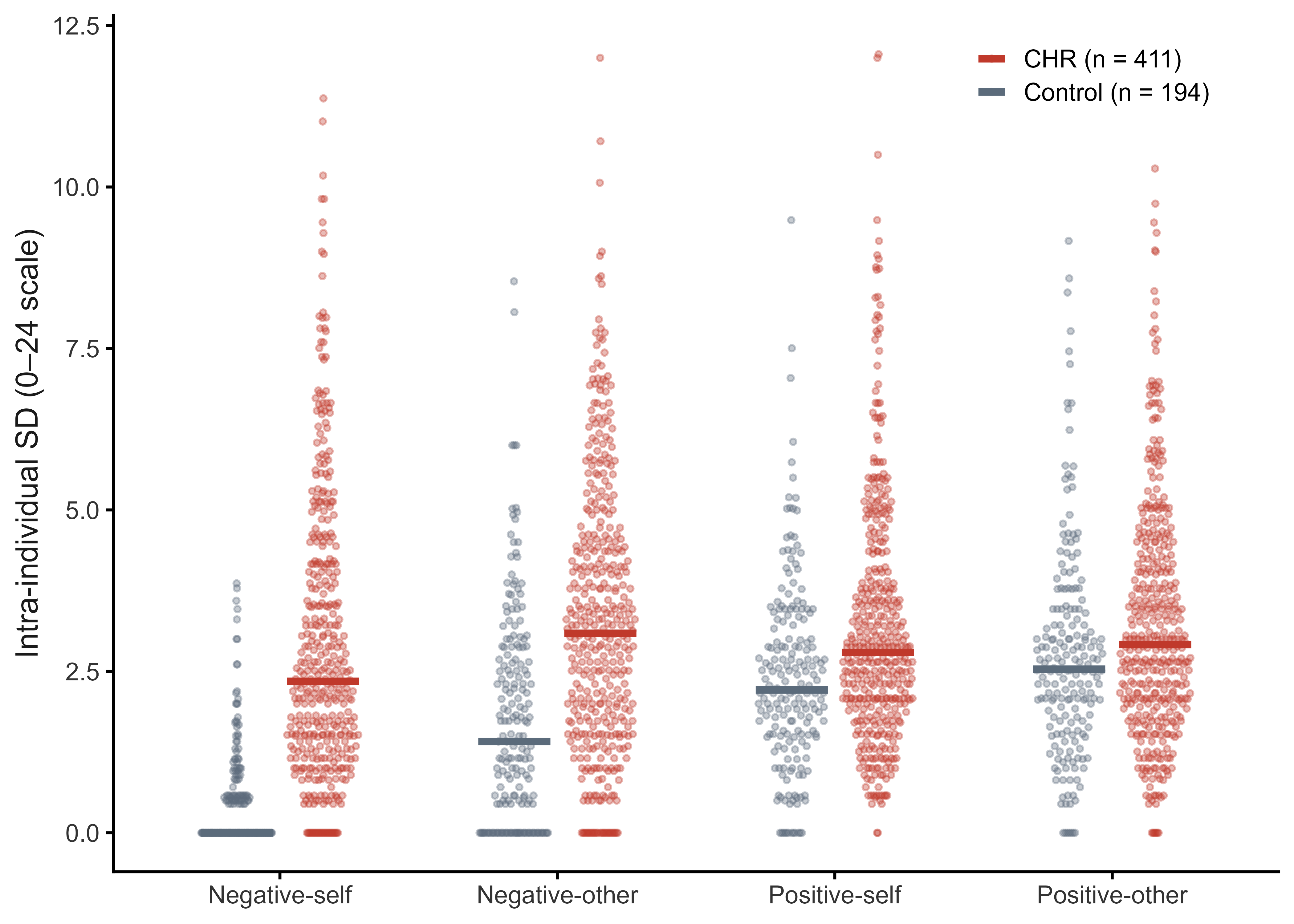

*Note.* Each point is one participant's intra-individual SD (iSD) across visits on a BCSS subscale (0–24 range), in the analytic sample (CHR *n* = 411, 410 for negative-other; control *n* = 194). Points are horizontally jittered by group within subscale; horizontal bars mark group medians. The control median on negative-self is 0.00 because 50.5% of controls scored 0 at every visit. BCSS = Brief Core Schema Scales; CHR = clinical high risk for psychosis.
